# The provision of information on time-lapse imaging: A systematic analysis of UK fertility clinics websites

**DOI:** 10.1101/2023.11.02.23297967

**Authors:** Manuela Perrotta, Letizia Zampino, Alina Geampana, Priya Bhide

## Abstract

**Research Question:** This study aims to systematically analyse the provision of information on Time-lapse Imaging (TLI) by UK fertility clinic websites.

**Design:** We conducted an analysis of 106 clinic websites that offer fertility treatment to self-funded patients. The analysis aimed to examine whether these clinics offer TLI, the associated cost for patients, and the clarity and quality of the provided information.

**Results:** Out of the 106 websites analysed, 71 (67%) claimed to offer TLI, with 17 being NHS clinics and 54 being private clinics. Among these websites, 25 (35.2%) mentioned charging patients between £300 and £850, 25 (35.8%) claimed not to charge patients, and 21 (29.6%) did not provide any cost information for TLI. Although TLI is generally considered safe for patients and embryos, only 21 (29.6%) websites provided information on the associated risks. Furthermore, 64 (90.1%) websites made claims or implied that TLI leads to improved clinical outcomes by enhancing embryo selection. Notably, 34 (47.9%) websites did not mention or provide any links to the HFEA traffic light system. Additionally, 30 (42.2%) websites made claims regarding the effectiveness of TLI that contradicted the assessment of the HFEA, referring to early, mostly unspecified, studies.

**Conclusions:** It is crucial to provide patients with clear and accurate information to enable them to make fully informed decisions about TLI, particularly when they are responsible for the associated costs. The findings of this study raise concerns about the reliability and accuracy of the information available on fertility clinic websites, which are typically the primary source of information for patients.

## Introduction

In recent years, a variety of additional tests, treatments and technologies – usually known as add-ons – have been introduced and offered to fertility patients on top of standard IVF/ICSI cycles. These novel fertility interventions have sparked heated professional, public and media debates due to the lack of evidence supporting their efficacy (Heneghan et al, 2016; Harper et al., 2017; Cochrane, 2021), the poor quality of information available (Spencer et al, 2016; Van de Wiel, 2020) and their potential mis-selling (CMA research, 2020).

In the UK, these concerns have prompted regulatory efforts by the Human Fertilisation and Embryology Authority (HFEA) and the Competition and Market Authority (CMA). The HFEA traffic light system, introduced in 2017, is a classification framework used in the UK to assess the safety and effectiveness of treatment add-ons (see HFEA, 2022a). It categorises treatments as green (safe and effective), amber (uncertain or limited evidence), or red (unsafe or ineffective). This system is meant to help guide patients and healthcare providers in making informed decisions about fertility treatment options. In a similar manner, in June 2021 the CMA published guidelines which included recommendations to enhance the quality and accessibility of information given to patients and avoid potential mis-selling of add-on treatments (CMA, 2021a and 2021b).

Particular attention has been paid to fertility clinic websites, as these are often the first point of information for patients (HFEA, 2019; CMA, 2020 and 2022a). The CMA carried out a review of information on add-ons available on clinics’ websites one year after the introduction of their guidelines for clinics (CMA, 2022b), showing compliance issues regarding the information provided about some of the add-ons under examinations. Concerns were raised about the lack of information on risks, insufficient clinical evidence, and misrepresentation of the HFEA traffic light system.

Notably, the CMA review did not cover time-lapse imaging (TLI), which according to a recent study (Van de Wiel et al., 2020) is the most common add-on offered by UK fertility clinics. The popularity of TLI is confirmed by two HFEA patient surveys, which indicate that this is the second most common add-on after acupuncture and its use in IVF cycles has increased from 19% in 2018 to 27% in 2021 (HFEA, 2019, 2022b).

To address this gap, this study investigates the provision of information on TLI through a systematic analysis of UK fertility clinic websites. Incorporating a camera inside the incubator, TLI allows continuous monitoring and recording of the development of embryos, providing valuable insights to fertility professionals – insights meant to aid in the selection of embryos most likely to lead to a successful pregnancy. Despite the various advantages TLI offers to professionals (see Perrotta and Geampana, 2020), at present TLI is amber on the HFEA traffic light system as there is no conclusive evidence showing that it can be effective in improving live birth rates. Although TLI is not considered a risk to patient or embryo health (HFEA, 2022a), concerns are raised regarding its high cost and whether it is acceptable to charge patients for using TLI without evidence that it increases their chances of having a baby (Armstrong et al., 2015 and 2019; Kieslinger et al., 2023).

With 66% of the cycles privately funded in 2019 (HFEA, 2021), patients must be provided with clear and accurate information to make fully informed decisions about the fertility treatment they are paying for. According to the CMA guidelines (2021), this should include information about costs, the potential add-on treatment benefits to the patient and, if relevant, any risks.

To conduct our analysis, we reviewed all the UK fertility clinic websites by collecting data on how TLI is presented in June 2022 – one year following the introduction of the CMA guidelines on how to present information to fertility patients. Drawing on these guidelines, we systematically analysed all the websites of UK fertility clinics. After a detailed description of the materials and methods, we present our analysis of how TLI is presented on clinics websites, its cost for patients, and the clarity and quality of information on TLI.

## Materials and methods

To identify all the fertility clinics in the UK offering TLI, we began referring to the HFEA (2021) list of licensed and active clinics for 2020/21, which consisted of 134 clinics (both NHS and private). We excluded 30 clinics that do not provide IVF/ICSI treatments (comprising 14 clinics dedicated solely to storage and 16 clinics dedicated solely to research). We identified 104 clinics in total: 44 NHS and 60 private clinics. During the process of locating the clinics’ websites, we further excluded 16 clinics (10 NHS and 6 private) as their individual websites were not found. We included an additional 24 websites of satellite clinics that are part of larger groups. These satellite clinics were considered separate entries due to variations within the groups in terms of TLI availability, cost, and presentation of information. As a result, we analysed all 106 identified clinic websites, comprising 34 NHS clinics and 72 private clinics.

Our analysis of the 106 clinic websites revealed that 71 websites (67%) claimed to offer TLI as part of their treatment options. Among these clinics, 17 were NHS clinics that also provided treatments to self-funded patients, while 54 were private clinics. It is worth noting that while TLI appeared to be quite prevalent among the NHS clinics we analysed, with 50% (17 out of 34) offering it, it seemed to be even more commonplace among private clinics, with 75% (54 out of 72) providing this service. It is important to mention that distinguishing between NHS and private clinics solely based on their websites was not always straightforward, and we had to rely on the official HFEA (2021) data when available. This information may not be immediately accessible to patients seeking relevant information.

**Figure 1.**
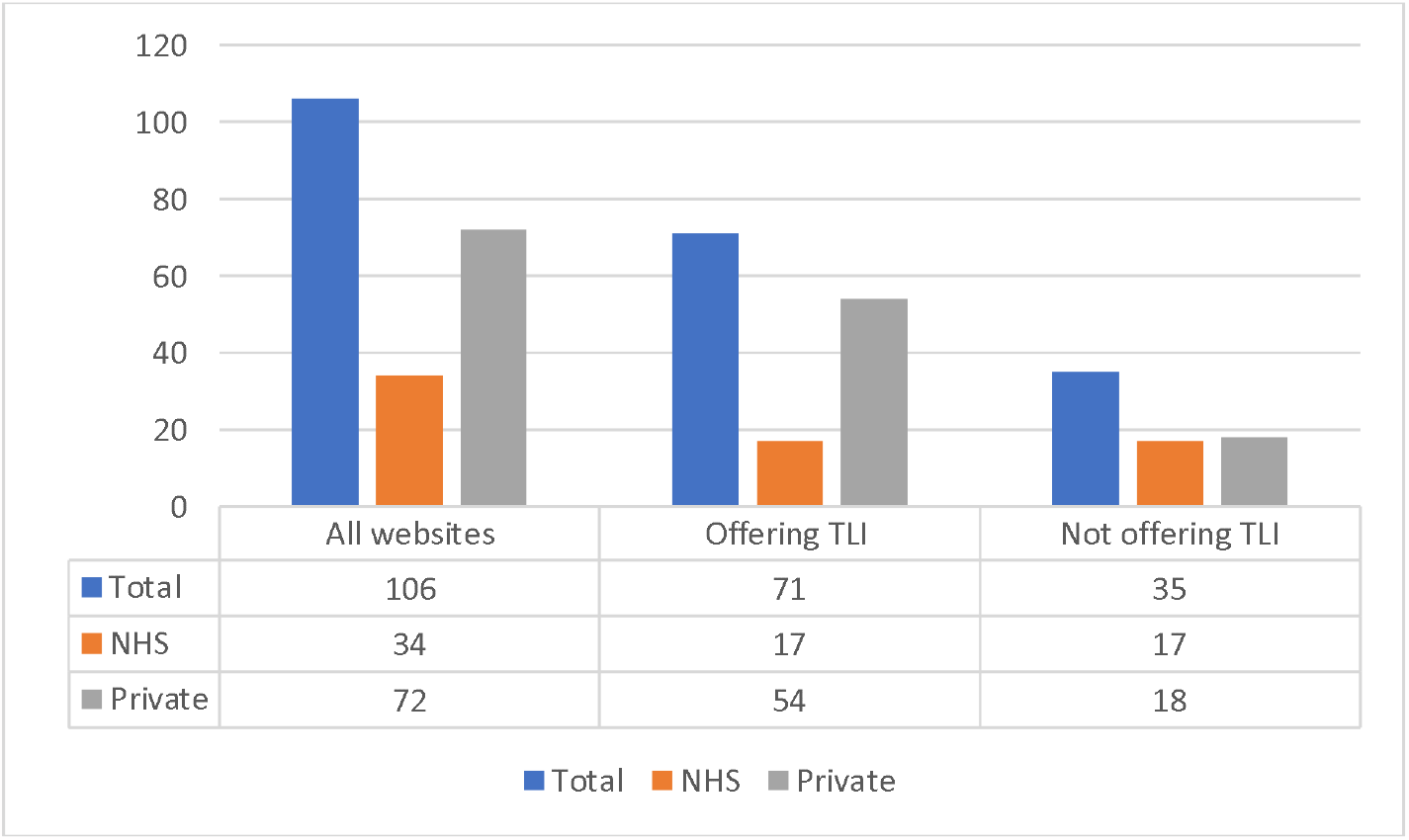
Number of clinics offering TLI

In June 2022, one of the authors (LZ) downloaded and saved as PDF files all webpages of these 71 websites containing information on TLI. These often included the main webpage if TLI was mentioned, a dedicated webpage to TLI, an additional webpage or separate PDF file with the pricelist, and any additional link or file related to TLI. Subsequently, all the information was anonymised and reported in a data matrix. In this matrix, a unique numeric code (from #1 to #71) was used instead of the name of the clinic. The data included in this matrix were organised under the following codes: 1. The description of TLI and any additional relevant information; 2. Information on the cost of TLI for patients; 3. Whether the website referred to the HFEA traffic light webpage on TLI and the text introducing the link; 4. Statements on the (potential) benefits of TLI; 5. Statements on the (potential) risks of TLI; 6. Statements on the evidence supporting the effectiveness of TLI. This data matrix was then reviewed by two additional authors (MP and AG) to reach an agreement on codes, for instance whether a certain statement was presented as a clear benefit of TLI or a generic description of the intervention.

When consensus on the data matrix was reached among researchers, further rounds of analysis were performed, comprising analyses of the content, cost and overall clarity and quality of information. For the latter, we draw on CMA guidelines included in their consumer law compliance review of fertility clinics on how consumer law applies to treatment add-ons:

> “Consumer law requires that existing and prospective patients are provided with material information at the time that they need it, and in a format that is clear and easy to understand. In our view this includes information about the risks, evidence base and the HFEA’s information about treatment add-ons, along with signposting to the HFEA’s website. This is so that the decisions patients make about whether to buy an add-on treatment are properly informed.” (CMA 2022b, p. 64)

In their review of fertility clinics, the CMA (2022b) also clarify that claims relating to the success rates of particular treatment add-ons should be accompanied by a clear explanation of the basis on which the claim is made, including what measure is being used (i.e., per embryo transfer or per cycle) and to which group of patients the success rate applies. The review emphasises that the absence or unclear presentation of this information can potentially mislead individuals regarding the advantages of a specific treatment add-ons, ultimately impacting their decision on whether to purchase them.

### How TLI is presented on clinic websites

In this first section, we present a content analysis of the wider narratives on how TLI is presented on clinic websites. As the CMA’s guidelines suggest that how information on addons is offered may influence patients’ decision to purchase TLI, in what follow we link any of the reported statements with the clinic websites, including information on the cost of TLI to patients (if available) or whether TLI is included in their standard package. As our interest is to analyse the provision of information on UK clinic websites as a whole rather than assessing how individual clinics provide information, we decided to anonymise the statements and refer to TLI or TLI-brand, when specific brands of TLI are mentioned.

Overall, the wider narratives on TLI seem to place emphasis on it as an advanced incubator technology and a tool that can potentially help professionals pick the best embryo, albeit without providing much detail on how this might be achieved in practice. Although this narrative most often suggests potential benefits of TLI in terms of clinical outputs, this is not always the case. In this section, we provide an analysis of the wider narratives used to present TLI, while we will focus on how clinic websites discuss the potential benefits of TLI in terms of clinical outcomes in the section on the quality of information.

Most fertility clinics describe TLI primarily as a cutting-edge incubator technology. The narrative presenting TLI as ground-breaking piece of laboratory equipment is very common and is found on both NHS and private websites, independently on whether clinics charge patients for TLI or not:

> “This state-of-the-art equipment allows our specialist laboratory team to closely monitor your embryo development in undisturbed conditions.” (NHS clinic #1, included)
>
> “We are proud to include TLI as standard for all embryology. TLI-brand is the world’s premier and highest-profile “time-lapse” embryo incubation and camera monitoring system. It captures detailed images of embryo development from the one cell zygote stage, shortly after fertilisation, right through to the fully expanded blastocyst.” (NHS clinic #3, included)

The potential advantage of undisturbed culture is emphasised along with TLI’s ability to take pictures of the developing embryos at regular intervals. As such, TLI is generally presented as a more ‘advanced’ type of incubator. In emphasising the undisturbed culture that a TLI incubator facilitates, clinics try to minimise patients’ fears about their embryos being disturbed in the lab. TLI is thus a technology that helps clinic staff protect the embryos from any potential outside interference. Some but not all websites go further in explaining the potential disadvantages of the traditional method of morphology observation where the embryologists take out embryos every day to study them briefly under the microscope. While this procedure is described as safe and established, clinics stress that TLI incubators remove the need for this practice due to their ability to monitor embryos from the inside with the help of the in-built cameras. TLI is presented as a technology that solves the potential problem of embryo conditions being disturbed by allowing staff to monitor them without taking them out of the incubator. Thus, the protective qualities of TLI are emphasised:

> “Designed to provide an individualised, undisturbed, optimal and stable environment, TLI gives each embryo the very best chance to develop, from fertilization through to embryo transfer. The incorporation of one of the most advanced time-lapse camera systems allows our embryologists to observe embryo development stage by stage. Whilst embryo safety is assured as each chamber is independently controlled and checked by a class leading monitoring system.” (private clinic #4, price £475)

Although risk minimisation is rarely mentioned explicitly when introducing TLI, the implication of describing TLI as a technology that facilitates undisturbed culture is that it offers less opportunities for the embryo’s optimal conditions to be disturbed. A minority of clinics explain incubator conditions as the ones that most closely resemble a women’s body. For instance, a website states that:

> “Standard in-vitro fertilisation takes place within the embryology laboratory following an egg collection. The embryologist will carefully combine the eggs and sperm within an environment designed to mimic the conditions of the womb. Fertilisation is allowed to take place naturally, with minimal intervention from the laboratory. Once the Embryologist has identified those eggs which have fertilised and become embryos, they will be moved into our TLI incubator to be monitored for 3-5 days”. (NHS clinic #69, price unclear)

Some websites do mention the capacity of TLI in terms of how many embryos would fit into one incubator. In addition, only a minority of websites describe in more detail the lab procedures around how and when embryos are placed in TLI. However, a few clinics do advise patients to talk to staff if they require more information on TLI.

Many descriptions of TLI emphasise the technology’s capacity for monitoring embryos, thus allowing embryologists to: 1) notice if the embryos have done something unusual or worrisome and 2) pick the best embryo based on the close study of morphology. Most websites emphasise the latter, however, without usually referring to any evidence that suggests that this is indeed the best way to pick the embryo that is most likely to result in a successful pregnancy and/or live birth (see the section on the quality of information). Nonetheless, TLI is generally presented as an advanced technology with a unique ability to facilitate the detailed study of embryo morphology:

> “The introduction of TLI allows us to monitor the developing embryos throughout the full course of their development without removing them from a stable incubated environment. The integrated camera and microscope automatically take an image of your embryos every 10 minutes to produce a time-lapse video of the vital stages of development enabling enhanced assessment by our team. This detailed development information allows us to identify only the best quality embryos for a future transfer procedure. (private clinic #35, price £500)

Only a minority of clinics mention that TLI is connected to computer software that also includes algorithms that can help embryologists score and classify embryos. Some of these websites mention the advantage of in-house algorithms developed using the clinic’s own TLI data.

Clinics tend to stress TLI’s ability to take pictures of embryos at regular intervals as a novel feature, especially when compared to traditional observation where it is only the embryologist that sees the embryo once a day under the microscope. Many, but not all clinics, also explain that the images taken can be put together to form a video where embryo development can be observed. Notably, only eight clinics mention on their websites that patients can be provided with a video of their implanted embryo. It is not clear based on the information provided when the video would become available.

Thus, although the visual capabilities of TLI are known, we have found that the technology is more likely to be presented as an advanced type of incubator rather than a visual technology that can offer patients insight into how their embryos are developing. The visual and monitoring advantages tend to be presented as something that staff need to make a potentially better selection decision rather than a souvenir or insight for patients.

### Cost analysis

We analysed the 71 websites identified to examine whether clinics include TLI in their standard package or charge patients who opt to have TLI in their treatment. It is worth noting that the range cost of basic treatment varies significantly across clinics, between £3,190 and £7,750, with an average of £4,380. As it is emphasised by the CMA review (2022a), a strict comparison of treatment prices is not possible, as clinics use very different way of presenting them and basic packages across clinics do not include the same elements. However, we signal that the average cost of treatment offered among the 17 NHS clinics that treat privately funded patients is £3,819, against an average of £4,547 among the 54 private clinic websites examined.

Of the 71 clinic websites analysed, 25 (35.2%) claim to charge a cost for TLI ranging between £300 and £850, and with an average of £614. Of these 25 clinics, 21 are private ones and four are NHS clinics. The latter charge patients for TLI respectively £450, £500 and £850 (two clinics). Adding complexity to the scenario, among the clinics that charge patients for TLI, nine offer special packages in which TLI is included. These packages include a combination of additional treatments, such as PGT-A, ICSI, or other various “advanced” laboratory techniques (as claimed on these websites).

Of the 46 remaining websites, 25 (35.2%) clearly state that TLI is included in the treatment and patients will not be charged for it. These include 11 NHS and 14 private clinics, with one of the NHS clinics specifying that TLI is available and included in the treatment but not guaranteed to all patients.

Despite stating that the clinic is equipped with TLI, the remaining 21 (29.6%) websites do not offer any information (including in their price lists) about the cost of TLI and it is not possible to determine from the websites whether TLI is included in their standard packages or not. These include two NHS clinics and 19 private ones.

**Figure 2.**
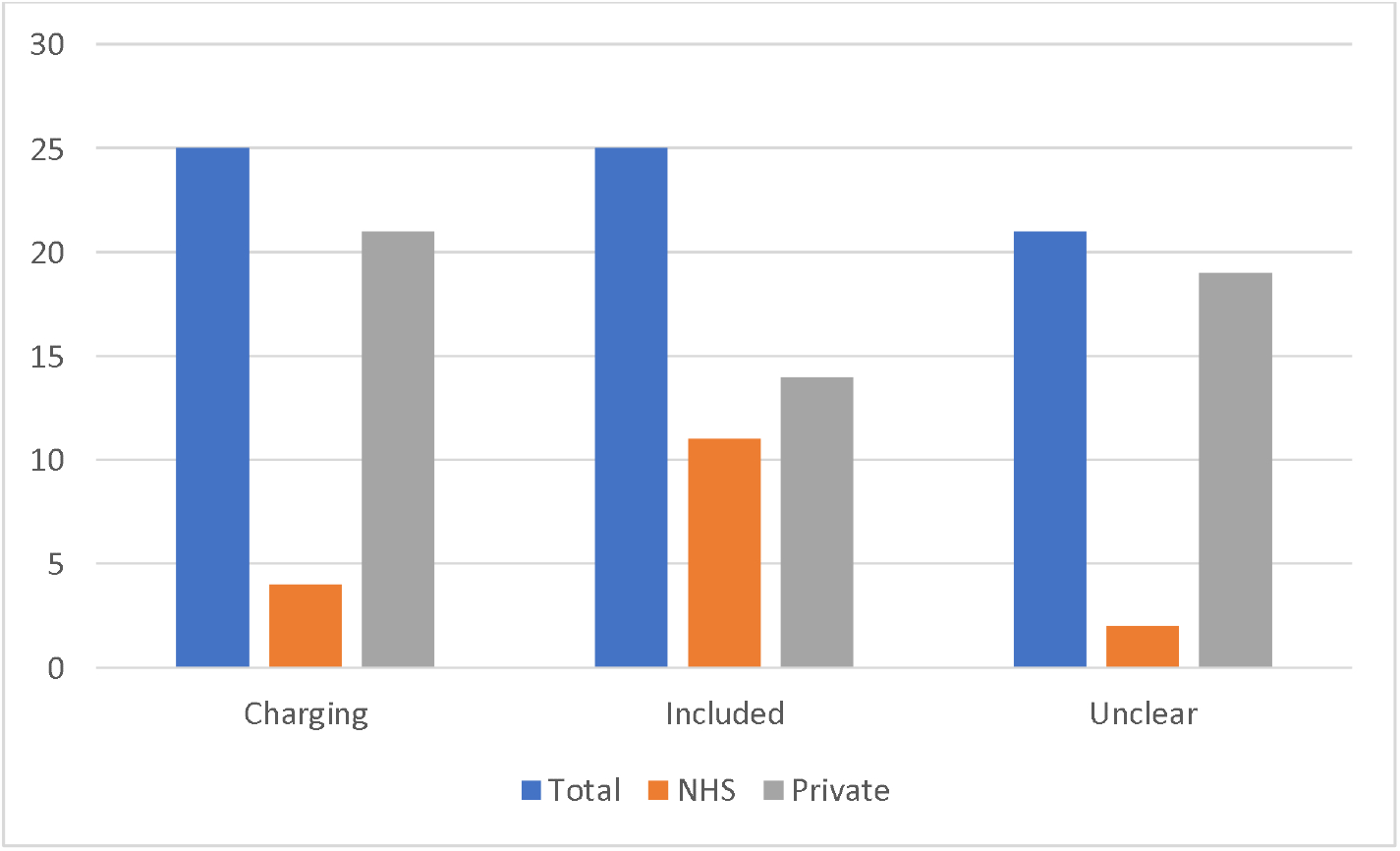
Information on TLI cost to patients

### Clarity and quality of information analysis

As pointed out above, our analysis of the clarity and quality of information offered by clinic websites follows the tenets of CMA’s guidelines on what information should be offered to patients. This includes information about the risks, benefits and the evidence base for these statements, including HFEA’s information about treatment add-ons.

#### How the risks of TLI are presented

According to the information available from the HFEA traffic light classification (2022a), “time-lapse imaging and incubation do not carry any additional known risks for the person undergoing fertility treatment or any child born as a result of fertility treatment”. Although the relevance of the information on risk might be less concerning than other add-ons, this remains important in terms of the clarity and quality of information offered on clinic websites.

Of the 71 websites analysed, only 21 (29.6%) offer information on the risks of TLI, including 5 NHS and 16 private clinics. The statements on the risks of TLI can be divided into two groups. 8 clinic websites report generic statements similar to the information of the HFEA: “there are no known risks to the woman or her embryos from TLI” (NHS clinic #38, included; private clinic #71, unclear)”. Interestingly, the remaining 13 clinic websites report an additional risk of TLI. For instance, one of these clinic websites states:

> “There are no risks that have been identified from the use of TLI. It is possible, however, that TLI might identify that none of your embryos are suitable for transfer and if that happens your doctor will work with you to identify the best way forward, taking account of all of the information from your treatment cycle, the embryo monitoring and your medical history.” (private clinic #25, price £850)

**Figure 3.**
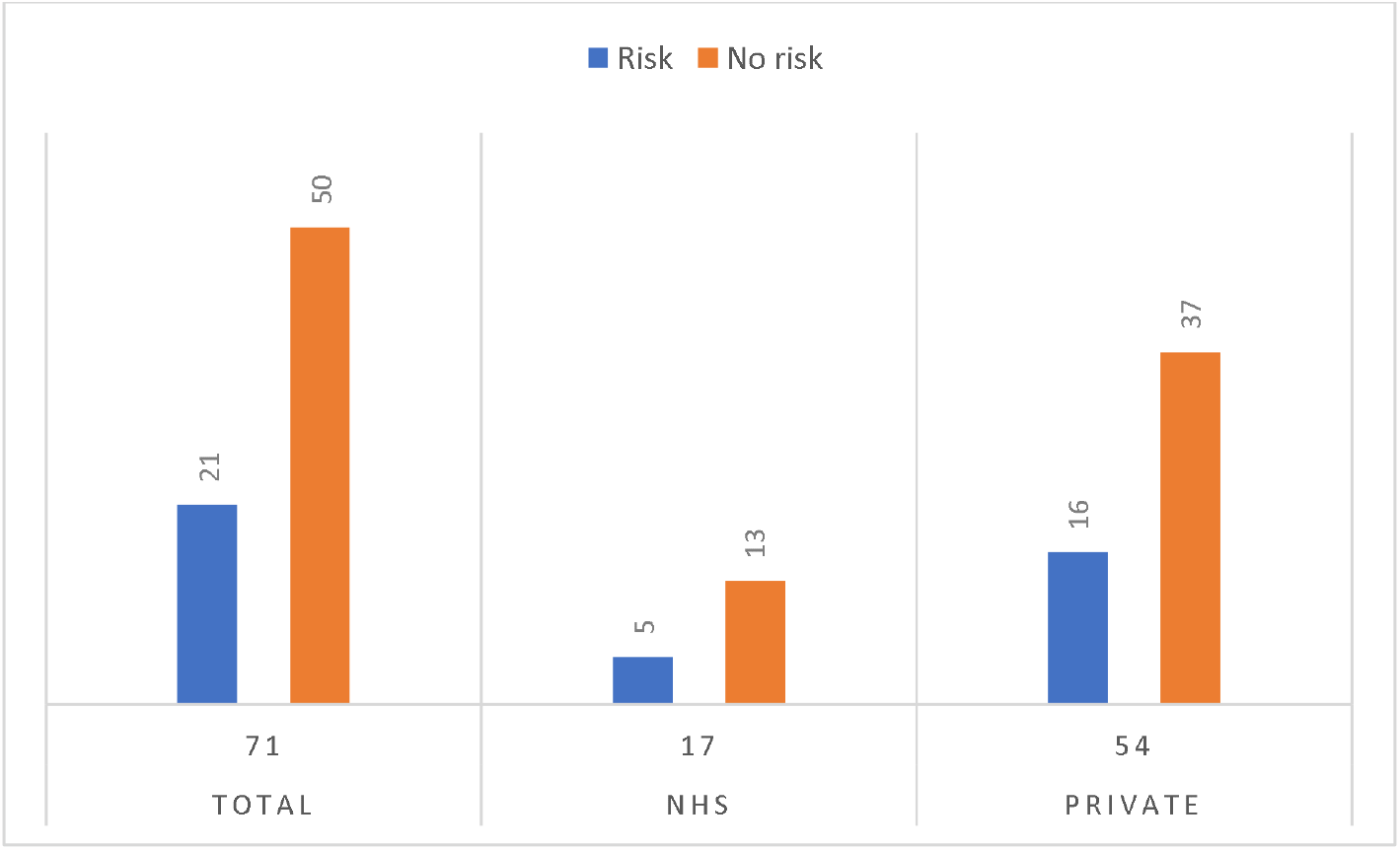
Statements on risks of TLI

#### How the benefits of TLI are presented

As mentioned above, the grand narrative on TLI emphasises a few key benefits, including the potential advantages of undisturbed culture and close monitoring, while less attention is paid to how TLI as a visual technology can offer patients insights into how their embryos are developing. Although these statements could potentially influence patients’ decision to purchase TLI as an add-on, they are not always indicating any specific benefit in terms of clinical outcome. In line with the aim of this article and CMA guidelines, in this section we focus specifically on the claims that, implicitly or explicitly, suggest that TLI can increase patients’ chances of having a baby.

Of the 71 websites analysed, only 7 (9.9%) clinic websites do not make any claim on TLI ability to increase chances of success. Three of these websites (one private and two NHS clinics) just mention they are equipped with TLI but do not have any information available on it. The other four websites (of a private and three NHS clinics) offer information on TLI without claiming any benefit in terms of clinical outcome.

The remaining 64 (90.1%) websites claim or imply improvements in clinical outcome due to the use of TLI. In what follows, we summarise the most common statements including some examples, while in the next section we discuss how the evidence base for these claims is presented. The total of these different statements is more than 64, as some of these websites make multiple claims. These claims vary significantly among clinics.

Three websites refer broadly to improvements in clinical outcome. For instance, among other advantages of TLI such as undisturbed culture or its ability to detect any abnormalities in cell division times and developmental behaviour, one of the websites claims that:

> “TLI-brand is the most widely adopted time-lapse system worldwide with documented improvements in clinical outcome”. (NHS clinic #12, price £500)

60 (84.5%) websites claim or imply an increase in clinical outcome due to TLI ability to support the selection of the best embryo(s), i.e. the embryos with the highest implantation potential or that are more likely to become a baby:

> “Enhanced information available to help identify the embryos with the highest potential for pregnancy”. (private clinic #28, included)
>
> “In IVF, TLI is used to help select the embryos most likely to successfully develop into a baby”. (NHS clinic #38, included)
>
> “With the increased information from TLI of embryo development and subsequent detailed analysis the embryologists can more confidently select the best embryos for transfer, significantly improving the chance of a successful single-embryo transfer. […] TLI can aid the IVF expert when assessing your developing embryos and help in selecting the best embryo(s) for transfer or freezing to optimise the chances of a successful pregnancy.” (private clinic #4, price £475)

**Figure 4.**
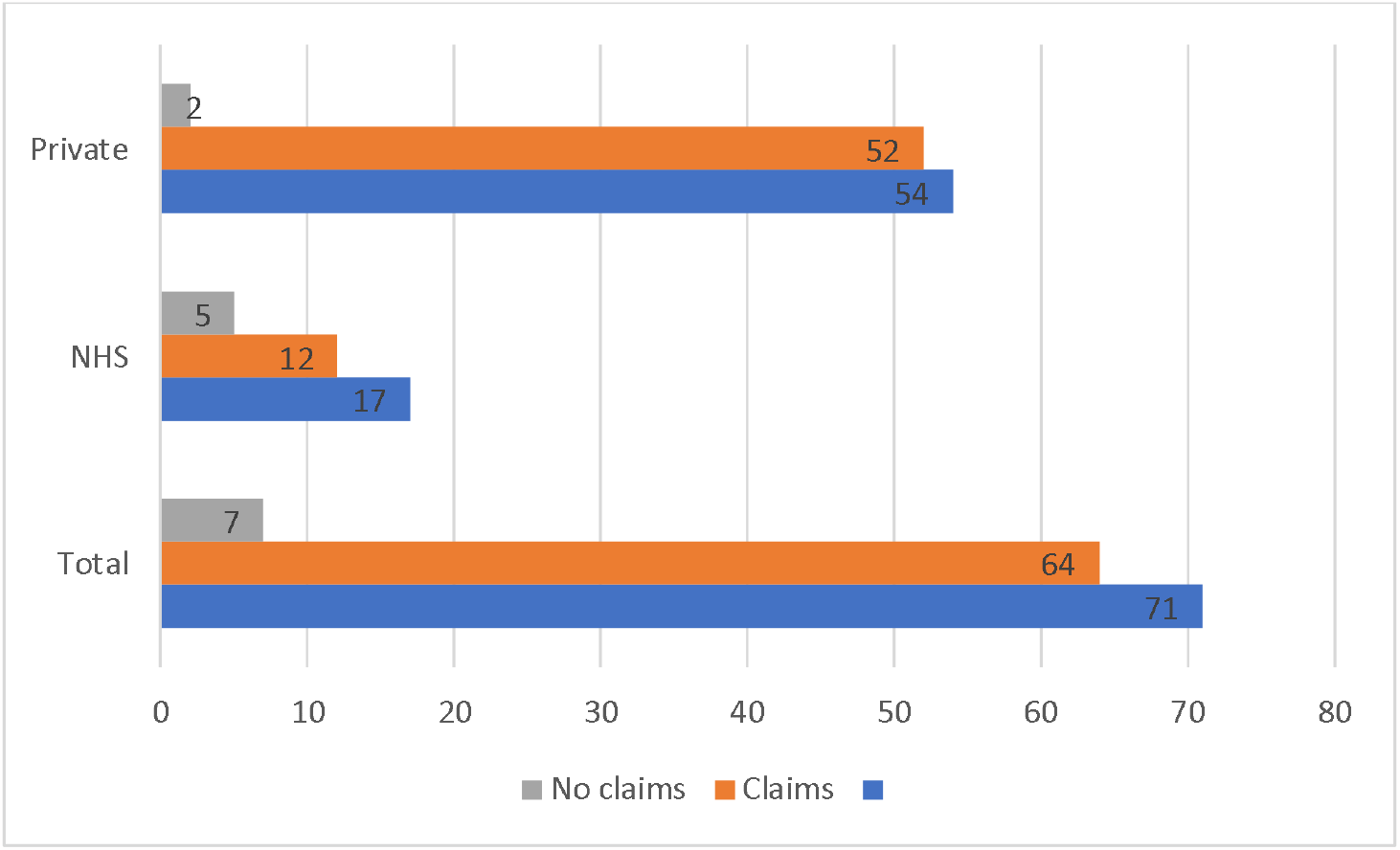
Claims on the benefits of TLI in terms of clinical outcome

Three websites suggest that TLI can improve the quality of the embryos:

> “Indeed, being undisturbed while they grow may improve the quality of the embryos”. (private clinic #71, price unclear)

Only one of the websites emphasises TLI ability to de-select embryos with potential issues:

> “It records the embryo development and allows us to analyse and compare the growth of each embryo, enabling us to select or exclude embryos for ET based on key information that is not available or apparent using traditional methods of embryo grading. Use of this technology allows us to avoid using embryos that have undergone abnormal development and would not be expected to implant or have a low chance of success”. (Private clinic #50, price unclear)

After explaining that, although TLI is used to help select the embryos most likely to successfully become a baby, the detailed information collected through TLI does not guarantee that an embryo will implant and result in a successful pregnancy and birth, one website claims that:

> “We have found that using TLI has also increased the proportion of patients who develop blastocysts and have embryos to freeze”. (NHS clinic #11, price £450)

While most of the clinic websites above do not specify whether the improvement in clinical outcome is for all patients or some groups, 27 (38%) websites offer specific information. For example:

> “Although in theory this technology can be applied to any type of patient undergoing IVF treatment, the chances of an improvement in the results are greatest among patients who generate more embryos because there is a better potential for selection. TLI is an embryo-selection tool that helps us more when we have a lot of embryos to choose from. It can be used in cases where more information about the embryo is desired in situations where there is repeated implantation failure, advanced maternal age and history of recurrent miscarriage. It will help couples and women to make an informed decision about future treatment plan or closure as appropriate. TLI will not be suitable for all patient groups. Please discuss this with your consultant if this technology can be useful to you. Until good quality RCT evidence is available, this technology is offered only in certain circumstances such as repeated implantation failure, after counselling as to its cost effectiveness. (private clinic #29, price unclear)

#### Evidence base and signposting to the HFEA’s website

In this last section, we presented the claims made on what are TLI benefits in terms of increasing clinical outcome. Most of the statements reported above did not report specific supporting evidence in the section of the websites where they were presented. A few referred to generic studies (without references) and some suggested the information was based upon the clinical experience of the team.

In this section, we discuss the evidence presented to justify the ability of TLI to increase success rates, focusing on whether or not these statements align with the assessment of the HFEA traffic light classification and whether the clinic websites signpost to the HFEA website.

34 (47.9%) of the 71 websites analysed do not have any signpost to the HFEA traffic light system. Of these 34, 10 are NHS clinics and 24 are private ones. Notably, none of the 7 clinics that do not make claims regarding the benefits of TLI in terms of clinical outcome include a signpost to the HFEA websites. 16 (22.5%) websites among those that include claims of TLI effectiveness do not present any alternative evidence supporting these claims. Among the remaining 11 (15.5%) websites, 10 private clinic websites mention non-specified studies suggesting that TLI is beneficial for some groups of patients. For example:

> “Studies suggest that embryos selected with the help of TLI have a high chance of forming a healthy pregnancy, so the technology will be especially welcome for patients with a poor reproductive record - that is, women who have already been unsuccessful in IVF and/or those of older reproductive age”. (private clinic #39, price £850).

In addition, one NHS clinic refers to a study (including a link to the company webpage where this study is available) of one of the companies producing and marketing a specific brand of TLI, which suggests that centres who use their product have better implantation rates:

> “Newly released data has compared UK centres with at least one (specific brand of) TLI to centres without a TLI. The data reveals an implantation rate (IR) in UK centres with a TLI as 38.3% compared to 30.5% in centres without a TLI. Implantation rate is defined as the number of gestational sacs observed at the 6 weeks pregnancy scan divided by the number of embryos that were transferred. The ‘uplift’ in IR of around 8% is similar to that reported in many of the published papers on the subject.” (NHS clinic #26, price £300)

Conversely, 37 (52.1%) clinic websites (7 NHS and 30 private) do include some signposting to the HFEA’s traffic light website. How the traffic light system is introduced by these websites varies significantly. These clinic websites either include short statements to refer to the traffic light system or a summary of its content:

> “For information from the HFEA on the risks and benefits of time lapse imaging click here.” (private clinic #28, included)
>
> “For more information on supplementary treatments, please visit the HFEA traffic light system on the HFEA website. The traffic light system gives further details on the most common treatment add-ons and how effective they are.” (NHS clinic #33, included)

**Figure 5.**
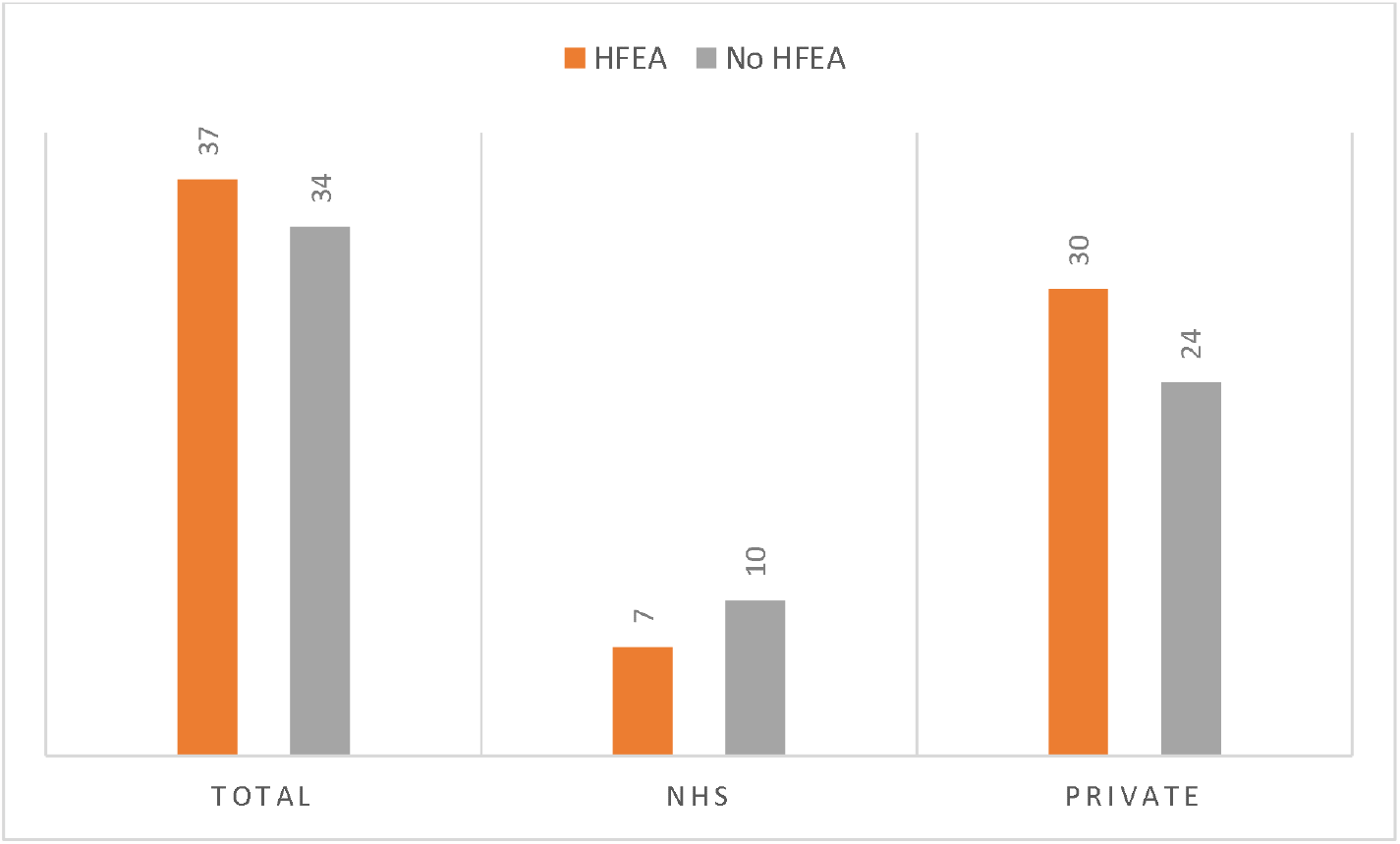
Websites signposting the HFEA traffic light system

References to the HFEA traffic light system are sometimes related to TLI specifically, while other times they refer to add-ons more generally. We report here some examples to illustrate the broad range of statements:

> “TLI is an optional additional treatment to routine IVF treatment, to ensure our patients make an informed decision about whether using TLI as part of their treatment the HFEA provide further information which can be found on their website here.” (private clinic #30, unclear)
>
> “Read further details below and for the latest on the effectiveness and safety of add-ons or adjuvants we recommend that you visit the HFEA website where our regulator has summarised the consensus of UK medical and scientific opinion.” (private clinic #5, price £300)
>
> “We support HFEA’s view on add-ons, please visit their webpage for more information.” (private clinic #66, price unclear)
>
> “It is important to remember that there are still relatively few robust research studies which show that Time Lapse Imaging will increase the chances of success. Please visit Treatment add-ons with limited evidence | Human Fertilisation and Embryology Authority for more information on treatment Add Ons. The Human Fertilisation and Embryology Authority (HFEA) are a government regulator, who ensure that fertility clinics and research centres comply with the law. The HFEA have provided TLI an amber rating. An amber rating means there is a conflicting amount of evidence on the effectiveness of this add-on treatment for improving your chances of having a baby. As a result, further research is still needed for this treatment. The HFEA reveals research into time-lapse imaging shows “promise” but is still too early to determine the effectiveness of this treatment. Initial data from studies support the idea that embryo selection or de-selection can be improved using TLI, and that embryo culture can be improved in an undisturbed environment. Both of these factors are important in improving the chance of success in IVF procedures”. (private clinic #4, price £475)

These websites clearly indicate that TLI is an add-on and correctly signpost to the HFEA websites. However, other sections of the websites often include claims regarding the ability of TLI to increase clinical outcome discussed in the previous section. 19 (26.8%) of these websites present additional information that conflicts with the assessment of the HFEA on TLI. For instance, among the clinics that present TLI benefits for specific groups, three private clinic websites include this additional claim – without any references – in the description of how TLI works:

> “Some retrospective studies have shown that embryos with specific division times and certain development patterns can have up to 15 – 20% better chance of pregnancy. The optimum times for cell division can be checked more easily and the chances of implantation improved in cases in which selection using TLI technology is possible.” (private clinic #29, price unclear)

Similarly, after a summary of the HFEA’s view on TLI, 13 clinics mention their own studies (without references or details) as evidence of the effectiveness of TLI for younger women due to their use of an in-house algorithm:

> “Our own data from a study published in 2017 of more than 23,000 treatment cycles showed a highly significant increase in births when TLI-algorithm was used to select embryos for patients aged younger than 38 using their own eggs. A paper published by us in 2019 showed that TLI-algorithm is superior for selecting embryos most likely to result in a birth than standard selection methods.” (private clinic #13, price £850).

Additional three websites include unsupported statements on TLI ability to significantly increase live birth rates or reduce miscarriages:

> “Early studies have shown an improvement in the chance of live birth by 56% over conventional methods of embryo selection.” (private clinic #50, price unclear)
>
> “A recent study showed a correlation between assessing the embryos via TLI and chromosomal integrity of the embryos. Choosing the embryos of low risk chromosomal abnormality improved the pregnancy rate by 56%. However, this is a preliminary small study and the conclusion needs to be confirmed in bigger prospective studies.” (private clinics #68, price £500).
>
> “Since TLI were introduced over eight years ago, over a million embryos have been cultured and there is now evidence that embryos cultured in a TLI will have a higher chance of implantation and a lower chance of early pregnancy loss”. (private clinics #52, price £450).

To summarise, 30 (42.2%) websites make claims on evidence supporting TLI that conflict with the assessment of the HFEA, referring to early, mostly unspecified, studies. These include three NHS and 27 private clinics. These claims are the only mention of evidence on 11 (15.5%) websites, while for the remaining 19 (26.8%) websites these claims are accompanied by a reference or a link to the HFEA traffic light website.

**Figure 6.**
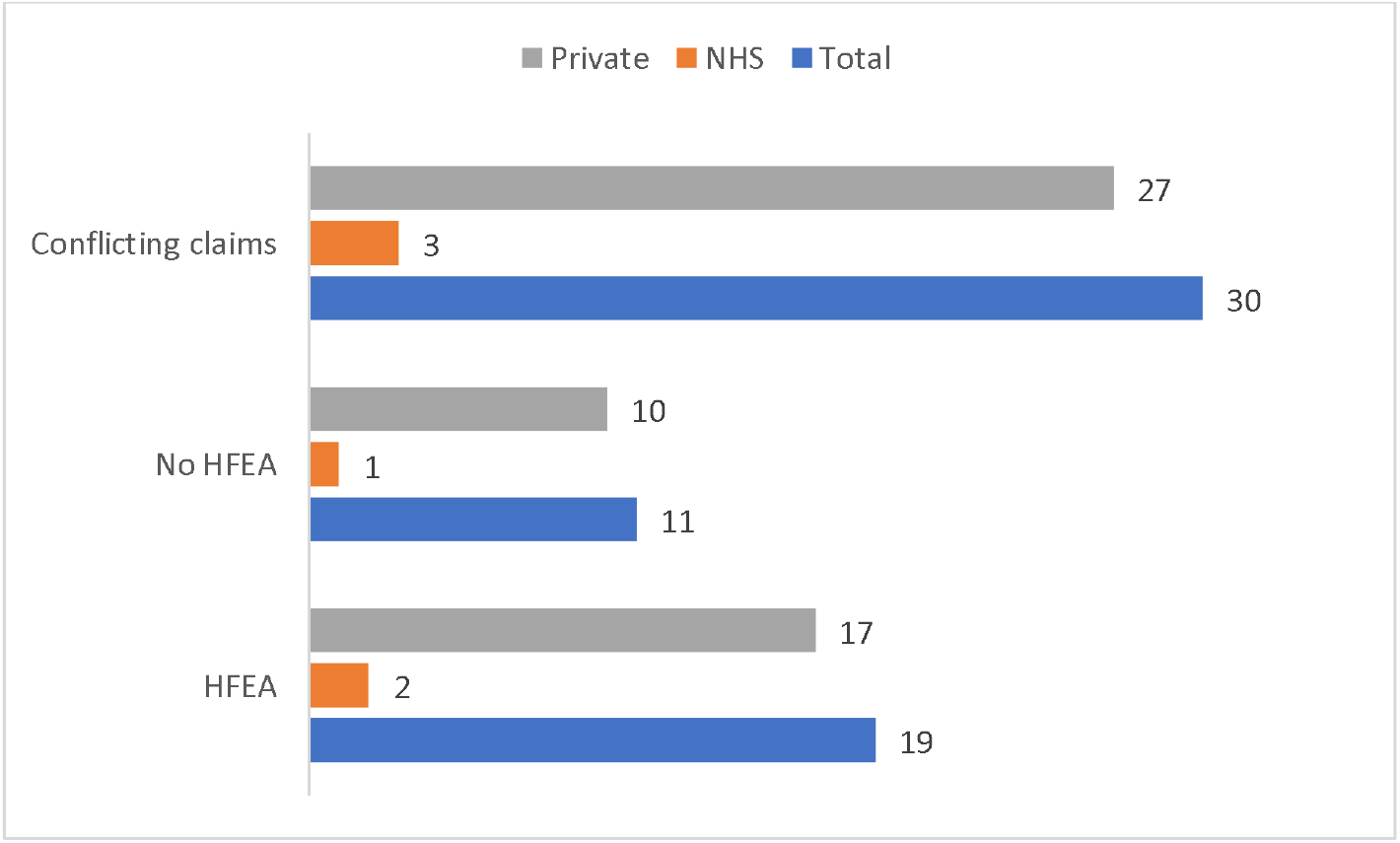
Claims on evidence supporting TLI in conflict with the HFEA traffic light

## Discussion

This study aimed to investigate the provision of information on TLI through a systematic analysis of UK fertility clinic websites. The findings shed light on several important aspects related to the availability, clarity, and quality of information regarding TLI on these websites. The findings reported in this article are consistent with the existing body of literature examining the quality of information provided on fertility clinic websites globally (Abusief et al., 2007; Sauerbrun-Cutler et al., 2021). This alignment is observed across both earlier studies (Spencer et al., 2016) and more recent investigations (Van de Wiel et al., 2020; Galiano et al., 2021; Lensen et al., 2021), which have specifically explored the provision of information concerning add-on treatments.

One of the key findings of this study is that a significant number of clinics (71 out of 106) claim to offer TLI on their websites. TLI has gained recognition and acceptance within the NHS setting, where a considerable proportion (50%) of clinics offered TLI as part of their services. Although TLI is quite prevalent in both settings, it appears to have become an integral part of the repertoire of services provided by private clinics, with a 75% offering this technology.

The provision of cost information for TLI on fertility clinic websites is an important aspect to consider when patients are making decisions about their treatments. However, the varying cost structures observed among the clinics raise concerns about financial transparency. Only over a third (35.2%) of the websites analysed clearly state that TLI is included in the treatment and patients would not be charged for it. Conversely, the same proportion of clinics (35.2%) charged patients a considerable fee to use TLI (between £300 and £850), indicating that the financial burden of TLI falls still largely on patients seeking treatment. Furthermore, while the large majority of websites (70.4%) provided information on the cost of TLI, a significant portion (29.6%) either did not disclose the cost or omitted this crucial information. This lack of transparency can significantly impact patients’ decision-making processes, particularly when they are self-funded and need to consider the financial implications of TLI. Moreover, the study reveals that less than a third (29.6%) of websites provide information on the potential risks associated with TLI. Although TLI is generally considered safe for patients and embryos, patients should have access to comprehensive and accurate information to make fully informed decisions about their fertility treatments.

A concerning finding of this study is the lack of mention or signpost to the HFEA traffic light system on a considerable number of websites (47.9%). Despite its limitations (Lensen et al., 2023), the traffic light system provides patients with an easily understandable rating system for the most common fertility treatment add-ons, including TLI. The absence of this information may hinder patients’ ability to make informed decisions. In addition, CMA guidelines make imperative for clinics to include a clear reference and link to the HFEA traffic light system to ensure transparency and facilitate patients’ access to crucial information.

More worryingly, most websites (90.1%) claimed or implied that TLI improves clinical outcomes by enhancing embryo selection. It is important to note that these claims are neither supported by the assessment conducted by the HFEA or the two available Cochrane reviews on TLI (Armstrong et al., 2015 and 2019). A significant percentage of websites (42.2%) claiming an increase on clinical outcome referenced early, unspecified studies that conflict with the HFEA’s evaluation. Only in few occasions links to these studies or refences are offered, but there is no clear discussion of what type of studies these are (mostly retrospective studies conducted by private companies). The discrepancy between the HFEA’s assessment and the conflicting evidence reported highlights the need for consistency and evidence-based claims on clinic websites. Patients rely on these websites as a primary source of information (HFEA, 2019; CMA, 2020 and 2022a), and it is crucial that the information presented aligns with the most up-to-date and reliable scientific evidence.

### Limitations of the study

This study has three key limitations. The first and most significant limitation pertains to the nature of the data collected. It should be noted that websites are dynamic entities, constantly subject to changes in content and pricing. Therefore, the analysis presented here reflects the information and prices available on clinic websites at a specific point in time (June 2022). This timeframe was chosen because it marked one year after the introduction of guidelines by the CMA, which clearly outlined the expected information regarding add-ons. Given the dynamic nature of websites, it is possible that some of the data discussed in this study may already be out of date. We hope that, if they have not done already, these findings will prompt clinics to promptly update and improve the clarity and quality of information on TLI available on their websites, taking into account the CMA guidelines.

The second limitation of this study is that only the information appearing on clinic websites was analysed. This means that information could be shared through additional advertising materials (such as leaflets or information sent to patients), open events hosted by clinics, and in consultations. While we acknowledge that these sources are also relevant for patients seeking information, it is worth noting that both the HFEA patient survey (2019) and the CMA research (2020, 20222) highlighted that clinic websites are a primary source of information for patients in the context of fertility treatment.

The third limitation pertains to the unavailability of data, either from the HFEA or other sources, regarding the usage of TLI by specific clinics and the number of cycles in which it is used. This limitation implies that we are unable to verify whether clinics claiming to offer TLI actually provide this service to their patients, or whether clinics that do not mention TLI on their websites may still offer it. The lack of data on TLI usage and availability poses a challenge in accurately assessing the actual provision of this technology by clinics.

These limitations should be taken into account when interpreting the findings of this study. Despite these limitations, the analysis of clinic websites provides valuable insights into the provision, transparency, and quality of information regarding TLI. Further research is warranted to address these limitations and obtain a more comprehensive understanding of the availability and usage of TLI across clinics.

## Conclusion

The case of TLI raises important considerations regarding the innovation model of fertility care. Challenges in generating reliable data on the effectiveness of add-on treatments (see Perrotta and Geampana, 2021) are often used as a justification to introduce interventions before robust evidence of their efficacy is available. This practice poses a particular concern in a sector where patients bear the financial burden of their treatment, essentially subsidising research into novel interventions. Previous research (Perrotta and Geampana, 2020) has highlighted various perceived advantages of TLI, including its utility as a laboratory tool, its potential for knowledge generation in embryology, and its role in managing patient expectations and treatment processes. This study confirms the widespread narrative presenting TLI as an advanced incubator and cutting-edge laboratory equipment. However, the justification for directly charging patients for the use of up-to-date technology remains unclear.

Furthermore, the increasing prevalence of TLI among clinics, particularly within the standard offerings of private clinics, may lead patients to assume that it significantly enhances their chances of achieving successful outcomes. The findings of this study, which examined the transparency and quality of information on TLI provided by UK fertility clinic websites, suggest that patients may be prone to overestimating the potential benefits of TLI for their treatment. These benefits, however, remain unproven, including the recent publication of a randomised controlled trial (Kieslinger et al., 2023) demonstrating that TLI, with or without the use of algorithms, does not improve patients’ chances of achieving successful pregnancies.

Previous studies (Perrotta and Hamper, 2021) have shown that patients often feel compelled to explore any available interventions that could enhance their chances of success in order to avoid future regret. Additionally, some patients are willing to embrace the experimental nature of fertility treatments and accept incremental risks when previous treatments have not yielded positive outcomes. However, many patients prefer to delegate the evaluation of evidence to medical professionals (Perrotta and Hamper, 2023) and rely on clinic websites as trusted sources of information (HFEA, 2019; CMA, 2020 and 2022a). Therefore, the information provided on clinic websites is crucial not only for individuals considering treatment at specific clinics but also for prospective and current patients seeking reliable information. The observed discrepancies in cost transparency, risk disclosure, claims on clinical outcomes, and adherence to regulatory guidelines raise concerns about the reliability and accuracy of information provided on these websites.

While the article highlights whether clinics charge for TLI or not, it is important to note that inaccurate information on clinic websites can be harmful to all prospective and current patients who heavily rely on the information provided by clinics and expect it to be trustworthy. Without clear and accurate information, patients are left without the necessary tools to make well-informed choices about their treatment. Therefore, fertility clinics should prioritise the enhancement of their websites to ensure the provision of accurate and evidence-based information, thereby empowering patients to make informed decisions regarding their fertility treatment.

## Data Availability

All data produced in the present study are available upon reasonable request to the authors

## Acknowledgements

The authors that the School of Business and Management at Queen Mary University of London for their support in data collection through a small grant.

## Author contribution statement

MP conceived the idea for the study and obtained a small grant from her institution to support data collection. LZ, AG, and PB were involved in the design of the study. LZ undertook data collection in June 2022. Analysis was undertaken by LZ, MP and AG. MP wrote the initial draft with the support of AG and LZ. All authors edited the first draft and approve the final version for submission.

## References

Abusief, M.E., Hornstein, M.D., Jain, T., American Society for Reproductive Medicine, Society for Assisted Reproductive Technology, 2007. Assessment of United States fertility clinic websites according to the American Society for Reproductive Medicine (ASRM)/Society for Assisted Reproductive Technology (SART) guidelines. Fertil Steril 87: 88–92.

S. Armstrong, N. Arroll, L.M. Cree, V. Jordan, C. Farquhar (2015), Time □lapse systems for embryo incubation and assessment in assisted reproduction, Cochrane Database Syst. Rev. (2)

Armstrong, S., Bhide, P., Jordan, V., Pacey, A., Marjoribanks, J., & Farquhar, C. (2019). Time □lapse systems for embryo incubation and assessment in assisted reproduction. Cochrane Database of Systematic Reviews, (5).

CMA (2020) Self-funded IVF Research: Qualitative Research Report. Available at: https://assets.publishing.service.gov.uk/media/5fa01b30e90e070420702a1b/IVF_Research_Final_Report.pdf

CMA (2021a) Fertility treatment: A guide for Clinics. Available at: https://www.gov.uk/government/publications/fertility-treatment-a-guide-for-clinics

CMA (2021b) Fertility treatment: A guide to your consumer rights. Available at: https://www.gov.uk/government/publications/fertility-treatment-a-guide-to-your-consumer-rights

CMA (2022a) Patients’ experiences of buying fertility treatment: Qualitative Research Report. Available at: https://assets.publishing.service.gov.uk/media/632c2fefe90e073721b08402/Consumer_research_report_160922.pdf

CMA (2022b) Consumer law compliance review of fertility clinics: Findings report. Available at: https://assets.publishing.service.gov.uk/media/632d65af8fa8f51d1f83391a/A._Final_findings_report.pdf

Cochrane (2021) Cochrane Special Collections: In vitro fertilisation – effectiveness of add-ons. Available at: https://www.cochranelibrary.com/collections/doi/SC000046/full

Galiano V, Orvieto R, Machtinger R, Nahum R, Garzia E, Sulpizio P, Marconi AM, Seidman D. “Add-Ons” for assisted reproductive technology: do patients get honest information from fertility clinics’ websites?. Reproductive Sciences. 2021 Dec;28(12):3466–72.

Harper, J., Jackson, E., Sermon, K., Aitken, R. J., Harbottle, S., Mocanu, E., …Lundin, K. (2017) Adjuncts in the IVF laboratory: Where is the evidence for ‘add-on’ interventions? Human Reproduction, 32(3), 485–491. 10.1093/humrep/dex004

Heneghan, C., Spencer, E. A., Bobrovitz, N., Collins, D. R. J., Nunan, D., Plüddemann, A., …Mahtani, K. R. (2016) Lack of evidence for interventions offered in UK fertility centres. BMJ, 355, i6295. 10.1136/bmj.i6295

HFEA (2019) Our national patient survey results. Available at: https://www.hfea.gov.uk/about-us/news-and-press-releases/2018-news-and-press-releases/our-national-patient-survey-results/

HFEA (2021), State of the fertility sector 2020/21. Available at: https://www.hfea.gov.uk/about-us/publications/research-and-data/state-of-the-fertility-sector-2020-2021/

HFEA (2022a) Treatment add-ons with limited evidence. Available at: https://www.hfea.gov.uk/treatments/treatment-add-ons/

HFEA (2022b) National Patient Survey 2021. Available at: https://www.hfea.gov.uk/about-us/publications/research-and-data/national-patient-survey-2021

Kieslinger, D. C., Vergouw, C. G., Ramos, L., Arends, B., Curfs, M. H. J. M., Slappendel, E., … & Lambalk, C. B. (2023). Clinical outcomes of uninterrupted embryo culture with or without time-lapse-based embryo selection versus interrupted standard culture (SelecTIMO): a three-armed, multicentre, double-blind, randomised controlled trial. The Lancet, 401(10386), 1438–1446.

Lensen S, Chen S, Goodman L, Rombauts L, Farquhar C, Hammarberg K. IVF add□ons in Australia and New Zealand: A systematic assessment of IVF clinic websites.Australian and New Zealand Journal of Obstetrics and Gynaecology. 2021 Jun;61(3):430–8.

Lensen, S., Armstrong, S., Vaughan, E., Caughey, L., Peate, M., Farquhar, C., … & Wainwright, E. (2023). “It all depends on why it’s red”: qualitative interviews exploring patient and professional views of a traffic light system for IVF add-ons. Reproduction and Fertility, 1(aop).

Perrotta M. & Geampana A. (2020), The trouble with IVF and randomised control trials: Professional legitimation narratives on time-lapse imaging and evidence-informed care, Social Science & Medicine, 258, 113115.

Perrotta M. & Geampana A. (2021), Enacting evidence-based medicine in fertility care: tensions between commercialisation and knowledge standardisation, Sociology of Health and Illness, 43(9), 2015–2030.

Perrotta M. & Hamper J.A. (2021), The crafting of hope: Contextualising add-ons in the treatment trajectories of IVF patients, Social Science & Medicine, Article 114317

Perrotta M. and Hamper J. (2023), Patient informed choice in the age of evidence-based medicine: IVF patients’ approaches to biomedical evidence and fertility treatment add-ons, Sociology of health and illness, 45(2), 225–241.

Sauerbrun-Cutler MT, Brown EC, Huber WJ, Has P, Frishman GN. Society for assisted reproductive technology advertising guidelines: How are member clinics doing?. Fertility and Sterility. 2021 Jan 1;115(1):104–9.

Spencer, E. A., Mahtani, K. R., Goldacre, B., and Heneghan, C. (2016) Claims for fertility interventions: A systematic assessment of statements on UK fertility centre websites. BMJ Open, 6, e013940. 10.1136/bmjopen-2016-013940

Van de Wiel, L., Wilkinson, J., Athanasiou, P. and Harper, J. (2020). The prevalence, promotion and pricing of three IVF add-ons on fertility clinic websites. Reproductive BioMedicine Online, 41(5), 801–806.

